# Genetically proxied therapeutic effect of metformin use, blood pressure and hypertension’s risk: A drug-target based Mendelian Randomization study

**DOI:** 10.1101/2023.04.12.23288119

**Authors:** Junhong Jiang, Di Hu, Qi Zhang, Zenan Lin, The μ- Biomedical Data Investigation Group (Mu-BioDig)

## Abstract

**AIMS:** Previous investigations on clarifying the metformin’s effect on blood pressure (BP) and hypertension provided inconsistent results. To evaluate the association of the genetically proxied effect of metformin drug targets on BP and risk of hypertension through a drug-target Mendelian Randomization (MR) analysis.

**METHODS:** 32 instrumental variables for five metformin targets (i.e., AMP-activated protein kinase (AMPK), growth differentiation factor 15 (GDF15), mitochondrial glycerol 3 (MG3), Mitochondrial complex I (MCI) and glucagon (GCG)) were introduced to the MR analysis on the outcome datasets of hypertension, systolic and diastolic blood pressure (SBP and DBP). The meta-analyses were conducted to acquire the general effect of the genetically proxied metformin use on hypertension risk and blood pressure.

**RESULTS:** The MR analyses demonstrated that the MCI- and MG3-specific metformin’s use would significantly reduce SBP, DBP, and the risk of hypertension. The meta-analyses showed that the genetically proxied metformin’s use equivalent to a 6.75 mmol/mol reduction on HbA1c could decrease both the SBP (Beta=-1.05, P <0.001) and DBP (Beta=-0.51, P=0.096). Furthermore, metformin’s use was also implied to reduce the risk of hypertension in two independent cohorts.

**CONCLUSIONS:** Metformin use may reduce blood pressure and hypertension risk. The MG3- and MCI-dependent metformin’s effect may play key roles in the anti-hypertension function.

## 1. Introduction

Hypertension is the leading modifiable risk factor for cardiovascular diseases, affecting more than 1.5 billion people globally.[1] Metformin, one of the most widely recommended first-line pharmacotherapy for managing type 2 diabetes (T2D), has been reported to improve cardiovascular outcomes which probably via lowering blood pressure (BP).[2] Fanghänel et al.[3] noted that metformin treatment lowered BP in patients with diabetes. Similar findings were reported in several randomized controlled trials (RCTs) involving patients with portal hypertension.[4] A meta-analysis of 26 RCTs containing 4119 patients indicated that metformin could effectively decrease systolic BP.[5] However, previous findings were inconsistent, with some studies indicating no remarkable effect of metformin on reducing BP.[5] Another meta-analysis based on 41 RCTs (3074 patients) reached the opposite conclusion that metformin had no significant effect on BP.[6] Notably, these studies had intrinsic methodologic limitations, including small sample sizes and selection bias. Whether metformin had an effect on BP and hypertension is currently unclear.

Mendelian Randomization (MR) analysis, a complementary and alternative approach to RCTs, is a powerful statistical method that uses the significantly associated single nucleotide polymorphisms (SNPs) as instrumental variables (IVs) to quantify potential causal effects. Drug exerts their effects by regulating the pharmacological targets’ expression, and the naturally occurring human genetic variation can serve as a proxy for therapeutic drug targets’ reaction.[7] A recently developed extension to the MR paradigm, i.e. the drug-target based MR study, has been used to find the drug-repurposing candidates for various diseases.[8] The objective of this study was to estimate the causal effect of metformin on hypertension in a larger European population using drug-target based MR. The present findings may potentially offer novel prevention and treatment strategies for hypertension.

## 2. Methods

### 2.1 The genetic instrumental variables (IVs) for five metformin targets

Zheng and colleagues have explored the causal relationship between metformin and Alzheimer’s disease risk through a drug-target based MR approach.[9] They have identified 32 instrumental variables for five primary drug targets (i.e., AMP-activated protein kinase (AMPK), growth differentiation factor 15 (GDF15), mitochondrial glycerol 3 (MG3), Mitochondrial complex I (MCI) and glucagon (GCG)) and 23 associated genes of metformin. (see Figure 1. and supplementary Table 7A of Zheng et al.[9]) In order to identify the valid IVs, their selection process not only complied with the three key assumptions of MR analysis but also performed additional tests. For instance, they confirmed the human tissue expression of the selected genes with the GTEX data, eQTLGen, and Zheng et al.[10-12] Besides, as a positive control, the authors verified metformin’s effect on reducing T2D risk and HbA1c using the MR method.[9] According to the authors, one standard deviation (SD) unit lowering of the HbA1c equals to 6.75 mmol/mol reduction of HbA1c. [9]

**Figure 1.**
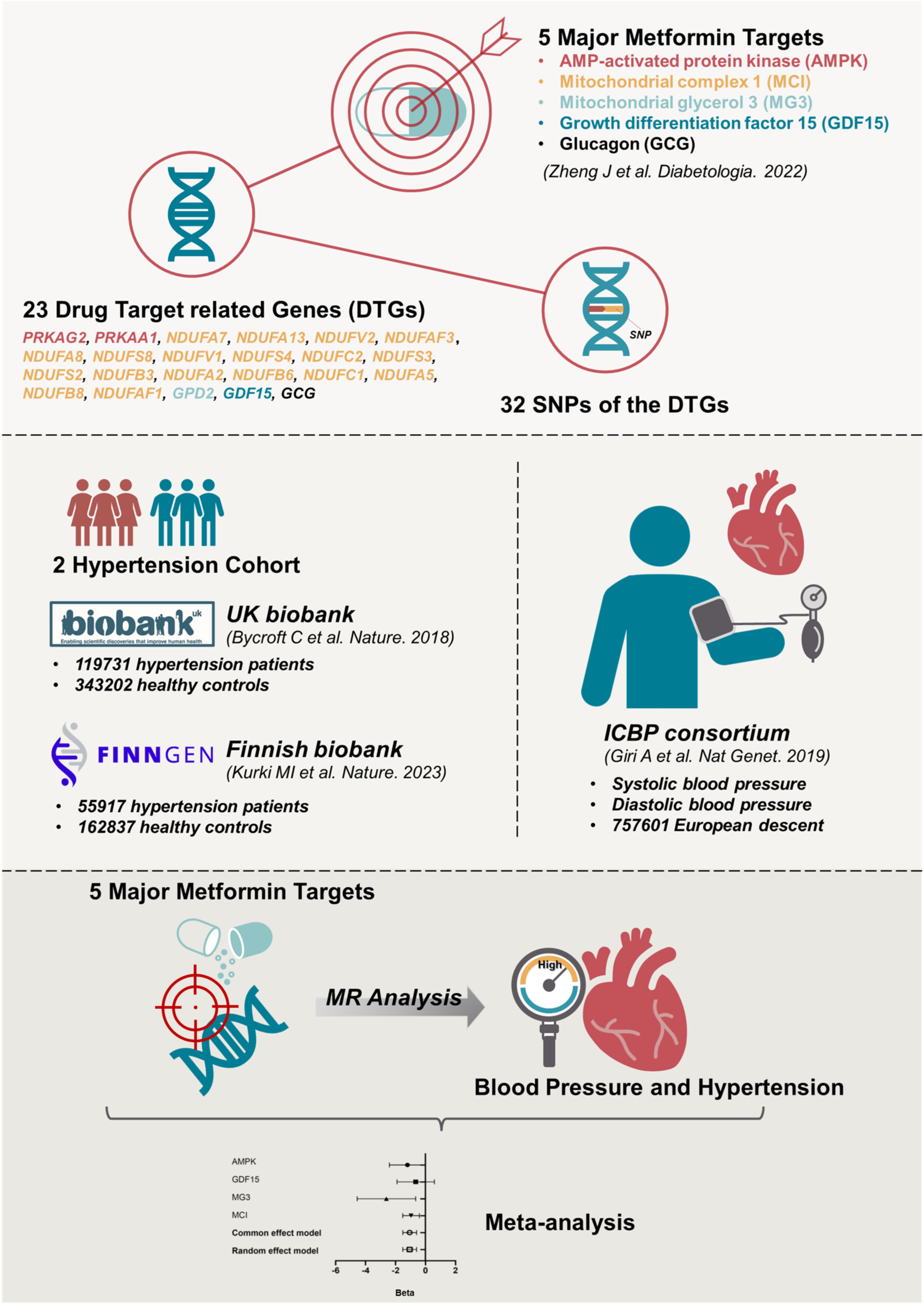
The workflow of the current work.

### 2.2 The information on the outcome datasets of hypertension and blood pressure

The GWAS datasets for hypertension were acquired from two independent cohorts. Hypertension cohort 1 came from the UK biobank and contained 462933 participants (119731 hypertension patients and 343202 healthy controls). Hypertension cohort 2 derived from the Finnish biobank project FinnGen (https://www.finngen.fi/fi). Its sample size was 218754, with 55917 being hypertension patients and 162837 being healthy controls. Their GWAS summary statistics data were acquired through the platform of the IEU OpenGWAS project (https://gwas.mrcieu.ac.uk/datasets, GWAS ID: “ukb-b-14057”, “finn-b-I9_HYPTENS”).[13, 14] The GWAS data for systolic blood pressure (SBP) and diastolic blood pressure (DBP) were provided by the International Consortium of Blood Pressure (ICBP).[15] The study included 757601 participants of European descent. The basic information of the included four outcome datasets was summarized in Figure 1.

### 2.3 MR analysis and meta-analysis

In the MR analysis, the inverse-variance weighted (when IVs>2) and Wald ratio (when IVs≤2) approaches were selected to be the major analytical tools. Other sensitivity methods such as MR Egger, Weighted median, Weighted mode and Simple mode were employed to assess the robustness of the conclusions of the MR analysis with IVs>2. The fixed-effect and random-effect statistical models were employed for the meta-analysis. All analyses were conducted with R packages MRPRESSO (version 1.0), ieugwasr (version 0.1.5), TwoSampleMR (version 0.5.6), and meta (version 6.0-0).[13, 14, 16] A *P* value less than 0.05 was considered statistically significant.

## 3. Results

### 3.1 The MR analyses of 5 drug targets on SBP, DBP and hypertension

The MR estimates of metformin drug targets on SBP, DBP, and 2 hypertension cohorts were summarized in Table 1. When the IVs for exposure and its potential proxy SNPs cannot be found in the outcome dataset, they were excluded from further analyses. Therefore, only four targets were kept for the analyses on SBP. We noticed that the MG3- and MCI-dependent metformin effects were confirmed to reduce both of SBP and DBP significantly. Consistently, the MCI-specific metformin influence was also identified to remarkably diminish the risk of hypertension in both tested hypertension cohorts. Besides, in hypertension cohort 1, the MG3- and AMPK-dependent metformin effects also demonstrated obvious anti-hypertension function. As shown in Table S1-S4, the pleiotropy tests of AMPK- and MCI-specific metformin’s effect on SBP, DBP and hypertension indicated no significant results. We also didn’t detect remarkable heterogeneity results in the AMPK study. However, the test of MCI demonstrated significant heterogeneity results. (See Table S1-S4) The sensitivity analyses on AMPK and MCI suggested that the effects were robust to various MR approaches. (see Table S5-S6)

### 3.2 Metformin’s effect on the SBP and DBP

In the MR analysis, the GCG-specific SNP and its proxy were not found in the GWAS dataset of SBP, and therefore, it was excluded from the meta-analysis. As shown in Figure 2A, the metformin’s targets could decrease both the SBP and DBP. In the analysis of SBP, four metformin targets were all confirmed to influence blood pressure reduction. The MG3-, MIC- and AMPK-specific metformin’s effects were statistically significant. Similarly, these four targets also demonstrated the ability to reduce the DBP, with the MG3- and MCI-specific effects being proven to be statistically significant. (see Figure 2B) Notably, the MG3-specific influence was confirmed to have the most substantial effect among the four targets.

**Figure 2.**
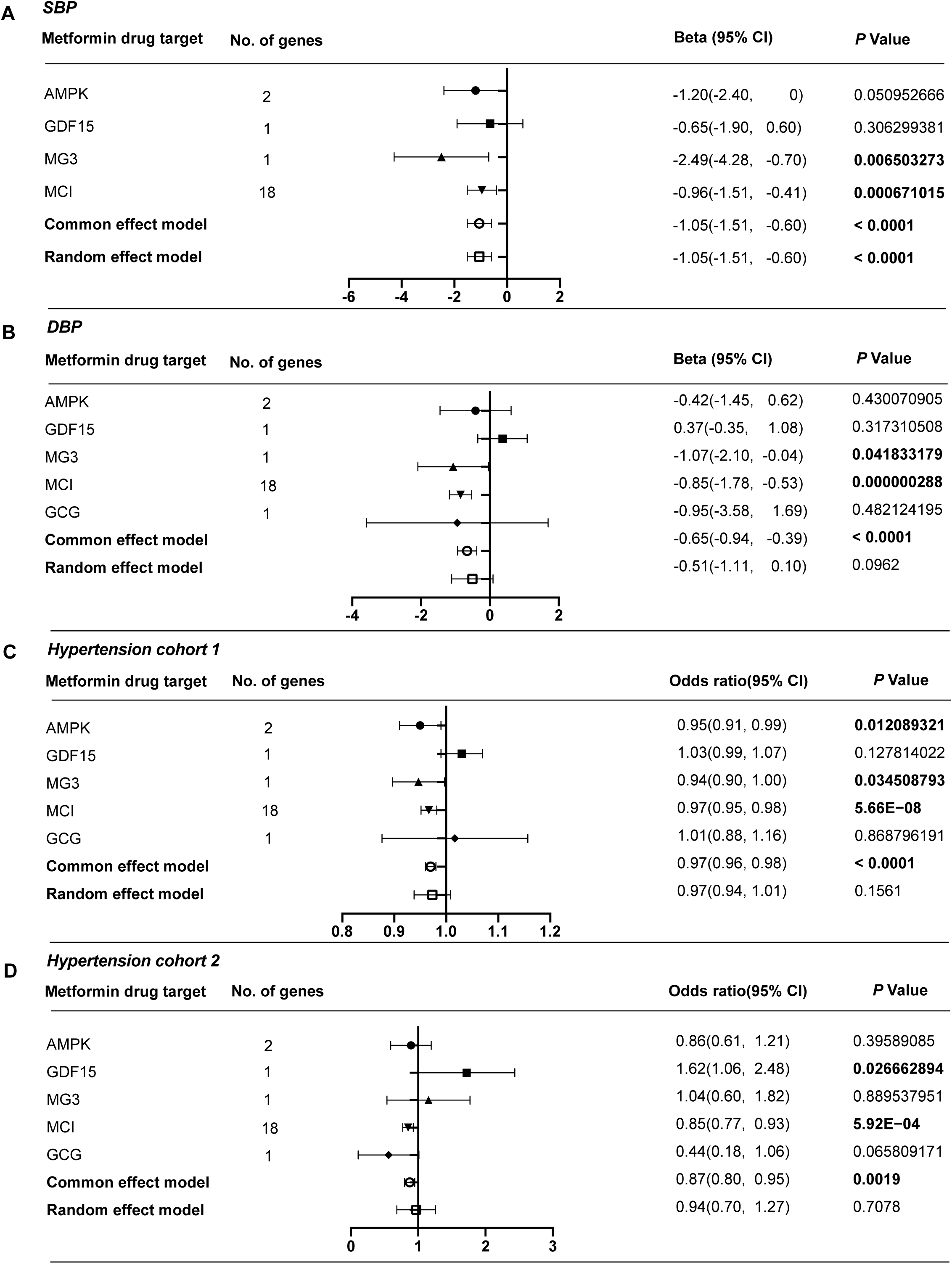
MR and meta-analysis of metformin’s effects on SBP (A), DBP (B) and two hypertension cohorts (C and D). The changes of SBP and DBP per 1-SD unit lowering of five targets-specific HbA1c level. The ORs of two hypertension cohorts per 1-SD unit reduction of five targets-specific HbA1c level.

### 3.3 Metformin’s effect on the risk of hypertension

In the MR analysis, we evaluated the effects of metformin on two independent hypertension cohorts. Though the random effect model meta-analysis suggested no remarkable effect of metformin target on reducing hypertensions’ s risk, the common effect model indicated a significant effect of metformin targets on decreasing hypertension risk (see Figure 2C and 2D.). Among five targets, the AMPK- and MCI-specific effects of metformin were consistent in two MR analyses and played key roles in reducing hypertension’s risk. In the MR analysis on hypertension cohort 2, the MG3-specific metformin’s effect showed no significant effect on the risk of hypertension. However, it was identified to reduce the risk of hypertension in cohort 1 significantly. The GCG-specific metformin’s effect was confirmed to have little effect on hypertension’s risk. The GDF15-specific metformin effect was implied to increase hypertension risk in two MR analyses.

## 4. Discussion

To the best of our knowledge, the present study is the first MR study to investigate metformin’s genetically predicted therapeutic effects on BP and hypertension. Basing on the data from large-scale GWAS studies in two independent hypertension cohorts of 175648 hypertensive patients, we observed that genetically proxied metformin use leads to a 13% reduction of hypertension risk, which may be owed to its ability to decrease SBP and DBP. Among five drug targets, metformin may exert its anti-blood pressure activities majorly through regulating MG3 and MCI. Collectively, these findings reported a suggestive benefit of metformin in reducing BP and provided novel evidence to guide hypertension prevention.

Metformin is recommended as the first-line treatment for T2D and is proposed to be beneficial for cardiovascular outcomes. Several earlier RCTs have suggested that metformin can significantly reduce BP in diabetic and hypertensive individuals. In addition to these human studies, several rodent studies have also provided evidence for BP-lowering effect of metformin. Kosegawa et al. observed that the SBP levels were significantly lower in the metformin group than in the controls group.[17] The effect of metformin on reducing BP in streptozotocin-induced diabetic rats was also observed in another animal trial conducted by Majithiya and Balaraman. [18] These results from RCTs and experimental studies are consistent with our findings. However, in contrast, some studies did not confirm a significant effect of metformin on reducing BP. Although the reason is unclear, the conflictive results might be due to multiple causes, such as small sample sizes, selection bias, differences in study populations, and strategies of BP measurement. Further prospective RCTs with multi-ethnic, larger sample sizes are needed.

Our findings suggest that MG3 and MCI may be the potential target for the effect of metformin on reducing BP. The MG3 target-associated gene *GPD2* encoded the mitochondrial glycerol 3-phosphate dehydrogenase (mGPDH), which is further partly used as a glycerol backbone for lipid molecules. Recent studies have shown that mGPDH was associated with kidney disease and macrophage inflammatory activities.[19, 20] We hypothesized that metformin might reduce BP through these mechanisms.[21] Furthermore, we also found that the inhibition of expression of an MCI-related gene could be associated with the effect of metformin on reducing BP. Intriguingly, the previous report has also confirmed that the inhibition of MCI would diminish hypertension in mice model.[22]

The current work still has limitations. First, though the 5 drug targets may recapitulate the major effect of metformin use, the possibility that some undiscovered targets would greatly affect the metformin response cannot be totally precluded. Second, future studies focusing on the validation of the included drug targets-associated genes (DTGs) are still needed since the present DTGs derive from the database and systemic literature review and thus lack the direct evidences from biological tests.[9] Third, our study analysed only the data from European participants which may limit the generalizability of the conclusion in other populations.

## 5. Conclusion

In summary, the present study provided novel evidences to support the causal effect of metformin on reducing hypertension risk. We revealed that metformin might decrease BP through the inhibition of MG3 and MCI. These findings implied the repurposing of metformin for cardiovascular diseases. However, randomized trials with large samples are still warranted to provide more robust conclusions.

## Data Availability

The used GWAS data were publicly available and their origins were described appropriately in the manuscript. The detailed information and codes required to reanalyse the data in this work are available from the corresponding authors upon reasonable request.

## Author Contributions

Conception, supervision and administration: Zenan Lin and Qi Zhang; Data curation: Junhong Jiang and Di Hu; Investigation: Junhong Jiang, and Di Hu; Methodology: Zenan Lin and Qi Zhang; Writing – original draft: Junhong Jiang and Di Hu; Writing – review & editing: Zenan Lin and Qi Zhang.

## Funding

No fund was obtained for this work.

## Conflict of Interest

The authors declare no conflict of interests.

## Ethics Approval

The used GWAS data were publicly available and approved by their corresponding institutions. An ethics approval for the current work is not required. No animal subjects were used in this work.

## Consent for publication

Not applicable.

## Acknowledgement

The authors want to express great thanks to the researchers and participants who made the relevant GWAS summary statistics publicly available.

---

**Table S1.** MR estimates of 5 targets-specific effects of metformin on SBP, DBP and hypertension.

| Outcome dataset | Exposure | Methods | No. of SNP | Beta | Standard error | P value |
| --- | --- | --- | --- | --- | --- | --- |
| SBP | AMPK | IVW | 3 | -1.196119026 | 0.612804128 | 0.050952666 |
| SBP | GDF15 | Wald ratio | 1 | -0.650518626 | 0.635881957 | 0.306299381 |
| SBP | MG3 | Wald ratio | 1 | -2.490827076 | 0.91531977 | **0.006503273** |
| SBP | MCI | IVW | 22 | -0.958757813 | 0.281891757 | **0.000671015** |
| DBP | AMPK | IVW | 3 | -0.41615111 | 0.527394197 | 0.430070905 |
| DBP | GDF15 | Wald ratio | 1 | 0.365916727 | 0.365916727 | 0.317310508 |
| DBP | MG3 | Wald ratio | 1 | -1.065242836 | 0.523415616 | **0.041833179** |
| DBP | MCI | IVW | 22 | -0.851825466 | 0.166006586 | **2.88E-07** |
| DBP | GCG | Wald ratio | 1 | -0.946154768 | 1.346091898 | 0.482124195 |
| Hypertension cohort 1 | AMPK | IVW | 3 | -0.054092043 | 0.021554683 | **0.012089321** |
| Hypertension cohort 1 | GDF15 | Wald ratio | 1 | 0.028217872 | 0.018530511 | 0.127814022 |
| Hypertension cohort 1 | MG3 | Wald ratio | 1 | -0.057142519 | 0.027029551 | **0.034508793** |
| Hypertension cohort 1 | MCI | IVW | 24 | -0.034164984 | 0.00629275 | **5.66E-08** |
| Hypertension cohort 1 | GCG | Wald ratio | 1 | 0.01145935 | 0.069371663 | 0.868796191 |
| Hypertension cohort 2 | AMPK | IVW | 3 | -0.147296363 | -0.147296363 | 0.39589085 |
| Hypertension cohort 2 | GDF15 | Wald ratio | 1 | 0.48301008 | 0.48301008 | **0.026662894** |
| Hypertension cohort 2 | MG3 | Wald ratio | 1 | 0.039453438 | 0.039453438 | 0.889537951 |
| Hypertension cohort 2 | MCI | IVW | 25 | -0.165961788 | -0.165961788 | **0.000591569** |
| Hypertension cohort 2 | GCG | Wald ratio | 1 | -0.821660719 | -0.821660719 | 0.065809171 |

## Notes

### Competing Interest Statement

The authors have declared no competing interest.

### Author Declarations

They have identified 32 instrumental variables for five primary drug targets (i.e., AMP-activated protein kinase (AMPK), growth differentiation factor 15 (GDF15), mitochondrial glycerol 3 (MG3), Mitochondrial complex I (MCI) and glucagon (GCG)) and 23 associated genes of metformin. (see Figure 1. and supplementary Table 7A of Zheng et al (Zheng et al. Diabetologia. 2022). The GWAS datasets for hypertension were acquired from two independent cohorts. Hypertension cohort 1 came from the UK biobank and contained 462933 participants (119731 hypertension patients and 343202 healthy controls). Hypertension cohort 2 derived from the Finnish biobank project FinnGen (https://www.finngen.fi/fi). Its sample size was 218754, with 55917 being hypertension patients and 162837 being healthy controls. Their GWAS summary statistics data were acquired through the platform of the IEU OpenGWAS project (https://gwas.mrcieu.ac.uk/datasets, GWAS ID: 'ukb-b-14057', 'finn-b-I9_HYPTENS').

